# GRADING prognostic factors for severe and recurrent *Clostridioides difficile* infection: expected and unexpected findings. A systematic review

**DOI:** 10.1101/2021.06.22.21259313

**Authors:** Tessel M. van Rossen, Rogier E. Ooijevaar, Christina M.J.E. Vandenbroucke-Grauls, Olaf M. Dekkers, Ed. J. Kuijper, Josbert J. Keller, Joffrey van Prehn

**Affiliations:** Amsterdam UMC, VU University Medical Center, Medical Microbiology & Infection Control, Amsterdam Infection & Immunity, Amsterdam, The Netherlands; Amsterdam UMC, VU University Medical Center, Gastroenterology & Hepatology, Amsterdam Gastroenterology Endocrinology Metabolism, Amsterdam, The Netherlands; Aarhus University, Clinical Epidemiology, Aarhus, Denmark; Leiden University Medical Center, Clinical Epidemiology, Leiden, The Netherlands; Leiden University Medical Center, Center for Infectious Diseases, Medical Microbiology, Leiden, The Netherlands; Haaglanden Medical Center, Gastroenterology & Hepatology, the Hague, The Netherlands; Leiden University Medical Center, Gastroenterology & Hepatology, Leiden, The Netherlands

## Abstract

**Background:** *Clostridioides difficile* infection (CDI), its subsequent recurrences (rCDI), and severe CDI (sCDI) provide a significant burden for both patients and the healthcare system. Treatment consists of oral antibiotics. Fidaxomicin, bezlotoxumab and fecal microbiota transplantion (FMT) reduce the number of recurrences compared to vancomycin, but are more costly. Identifying patients diagnosed with initial CDI who are at increased risk of developing sCDI/rCDI could lead to more cost-effective therapeutic choices.

**Objectives:** In this systematic review we aimed to identify clinical prognostic factors associated with an increased risk of developing sCDI or rCDI.

**Methods:** PubMed, Embase, Emcare, Web of Science and COCHRANE Library databases were searched from database inception through March, 2021. Study selection was performed by two independent reviewers on the basis of predefined selection criteria; conflicts were resolved by consensus. Cohort and case-control studies providing an analysis of clinical or laboratory data to predict sCDI/rCDI in patients ≥18 years diagnosed with CDI, were included. Risk of bias was assessed with the Quality in Prognostic Research (QUIPS) tool and the quality of evidence by the Grading of Recommendations Assessment, Development and Evaluation (GRADE) tool, modified for prognostic studies. Overview tables of prognostic factors were constructed to assess the number of studies and the respective direction of an association (positive, negative, or no association).

**Results and conclusions:** 136 studies were included for final analysis. Higher age and the presence of multiple comorbidities were prognostic factors for sCDI. Identified risk factors for rCDI were higher age, healthcare-associated CDI, prior hospitalization, PPIs started during/after CDI diagnosis and previous rCDI. Some variables that were found as risk factors for sCDI/rCDI in previous reviews were not confirmed in the current review, which can be attributed to differences in methodology. Risk stratification for sCDI/rCDI may contribute to a more personalized and optimal treatment for patients with CDI.

## Background

*Clostridioides difficile* infection (CDI), its subsequent recurrences (rCDI), and severe CDI (sCDI) provide a significant burden for both patients and the healthcare system(1). Antibiotic treatment with oral vancomycin and fidaxomicin are the cornerstone of CDI treatment(2-4). Fidaxomicin reduces the number of recurrences compared to vancomycin (5). Bezlotoxumab, a monoclonal anti-Toxin B antibody, can be added to oral anti-CDI antibiotic therapy and reduces the number or recurrences in patients at high risk to develop rCDI (6). Fecal microbiota transplantation (FMT) is a highly efficacious treatment for rCDI, preventing subsequent recurrences, and can lower mortality risk and disease burden of sCDI (7, 8).

The costs of fidaxomicin, bezlotoxumab and FMT are higher compared to those of vancomycin. Identifying patients diagnosed with initial CDI who are at an increased risk of developing a severe episode or recurrences could lead to more cost-effective therapeutic choices. Identifying patients at risk, however, is challenging. Several prediction models have been developed (9-30), yet none has been widely adopted in clinical practice.

### Objectives

In this systematic review we aimed to identify clinical prognostic factors associated with an increased risk of developing sCDI or rCDI.

## Methods

### Data sources

Two searches were performed to identify risk factors for (1) sCDI and (2) rCDI. The search terms and strategy were constructed by a trained librarian, see **Supplementary data**. PubMed, Embase, Emcare, Web of Science and COCHRANE Library databases were searched on October 4^th^, 2019. An update of the search was performed on March 13^th^, 2021. The search was restricted to articles published in English. Meeting abstracts were not considered. The search was not limited by year of publication. References of key papers were assessed for relevant papers.

### Study eligibility criteria

Study eligibility was assessed in a two-step selection process. Two independent reviewers screened title and abstract for potentially eligible articles (TvR and RO); discrepancies were resolved by consensus. Full-text articles were retrieved for eligibility; data extraction and risk of bias assessment of included studies was performed by two researchers (TvR and RO).

Inclusion criteria: prospective and retrospective cohort and case-control studies including patients ≥18 years, diagnosed with CDI, and providing an analysis of clinical or laboratory data to predict severe/recurrent CDI.

Exclusion criteria: study in specific patient populations with a distinct medical condition other than CDI (e.g. study population comprises only hemodialysis patients), and small studies (less than 30 patients with severe/recurrent CDI). Laboratory values that were studied as prognostic factors, but which are not part of the regular work-up in CDI (e.g. assays for specific cytokines). Variables that are part of (some of) the definitions of sCDI (such as leukocytosis) were excluded as prognostic factor for sCDI to avoid circularity.

Studies on prognostic factors with conflicting results in the uni- and multivariable or with contradicting results when compared to previous meta-analyses, were assessed and graded by a third researcher (CV-G and JK) and scrutinized by weighing the results by the quality of evidence per study.

Outcomes of interest were:

1. Severe CDI:
  a. ESCMID 2014/2021 definition: an episode of CDI with (one or more specific signs and symptoms of) severe colitis or a complicated course of disease, with significant systemic toxin effects and shock, resulting in need for ICU admission, colectomy or death (3, 31, 32); or:
  b. IDSA/SHEA definition: an episode of CDI with a white blood cell count of ≥15×10^9 cells/mL or a serum creatinine level >1.5 mg/dL or increase of 50% or greater from baseline(33); or:
  c. Any other author-constructed definition of severe/complicated/fulminant/fatal CDI.
2. Recurrent CDI: Recurrence is a new episode of CDI within 2-12 weeks after a previous episode, provided the symptoms of the previous episode resolved after completion of initial treatment (34, 35).

### Assessment of risk of bias

Risk of bias was assessed with the Quality in Prognostic Research (QUIPS) tool (36), which is recommended by the Cochrane Prognosis Methods Group. The QUIPS tool appraises six domains: (1) study participation, (2) study attrition, (3) prognostic factor measurement, (4) outcome measurement, (5) study confounding and (6) statistical analysis and reporting. The overall risk of bias per study is scored as low, moderate, or high.

### Quality assessment and data synthesis

The quality of evidence per study was assessed by using the Grading of Recommendations Assessment, Development and Evaluation (GRADE) tool, modified for prognostic studies (**Figure 1**) (37). This tool is recommended by the Cochrane Prognosis Methods Group and consists of eight domains. The starting point for the quality of evidence is based on the phase of investigation. The quality can be up- or downgraded according to seven other domains. The QUIPS score is included in the second domain (‘Study limitations/risk of bias’) of the GRADE tool. The outcome of this assessment is the quality of evidence per study, which can be very low (+), low (++), moderate (+++) or high (++++).

**Figure 1.**
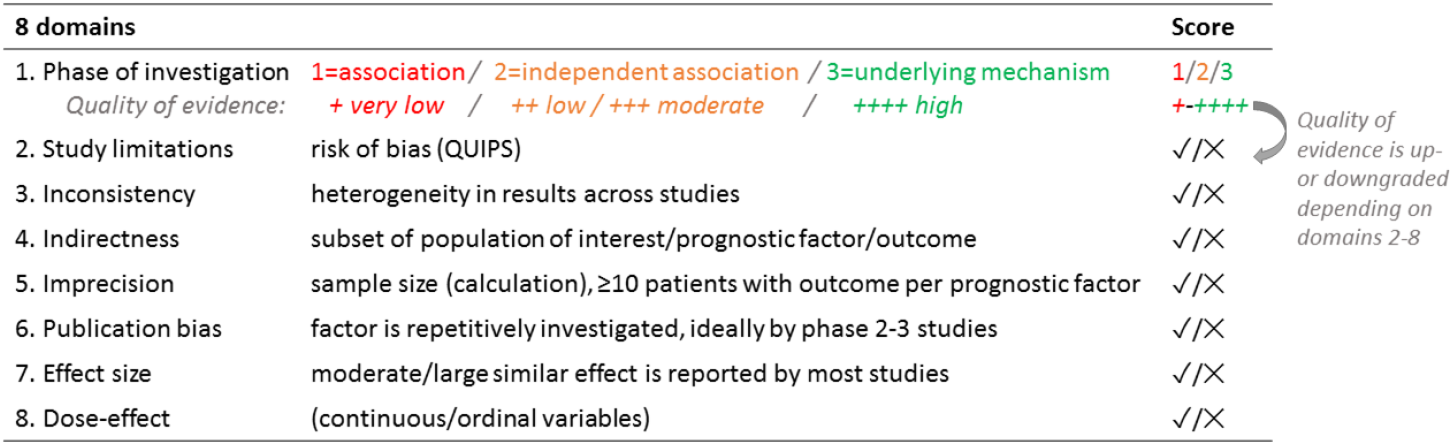
Grading of Recommendations Assessment, Development and Evaluation (GRADE) tool, modified for prognostic studies. The final outcome is the overall quality of evidence per study, which can be very low (+), low (++), moderate (+++) or high (++++).

Overview tables of prognostic factors were constructed to assess the number of studies and the respective direction of an association, i.e. positive, negative, or no association based on effect estimates, 95% CI intervals and p values, stratified by univariate and multivariate analyses. Similar prognostic factors (e.g. coronary artery disease and myocardial infarction) were combined to one factor for a more compact overview. No formal meta-analyses were performed.

## Results

The search for prognostic factors for sCDI yielded 1242 references; 126 studies were assessed in more detail and 76 were included for analysis (20, 24, 25, 29, 38-109); twelve more studies retrieved from cross-references were also included (110-121) resulting in 88 studies for final analysis **(Figure 2)**. The search for prognostic factors for rCDI yielded 1104 references; 105 studies were assessed in more detail and 36 were included for analysis (9, 10, 12-15, 102, 109, 122-149) as were seven studies from cross-references.) (111, 150-155). This resulted in 43 studies for final analysis **(Figure 2)**. Additionally, data of the pivotal RCTs on fidaxomicin and bezlotoxumab were used to support findings (156-160).

**Figure 2.**
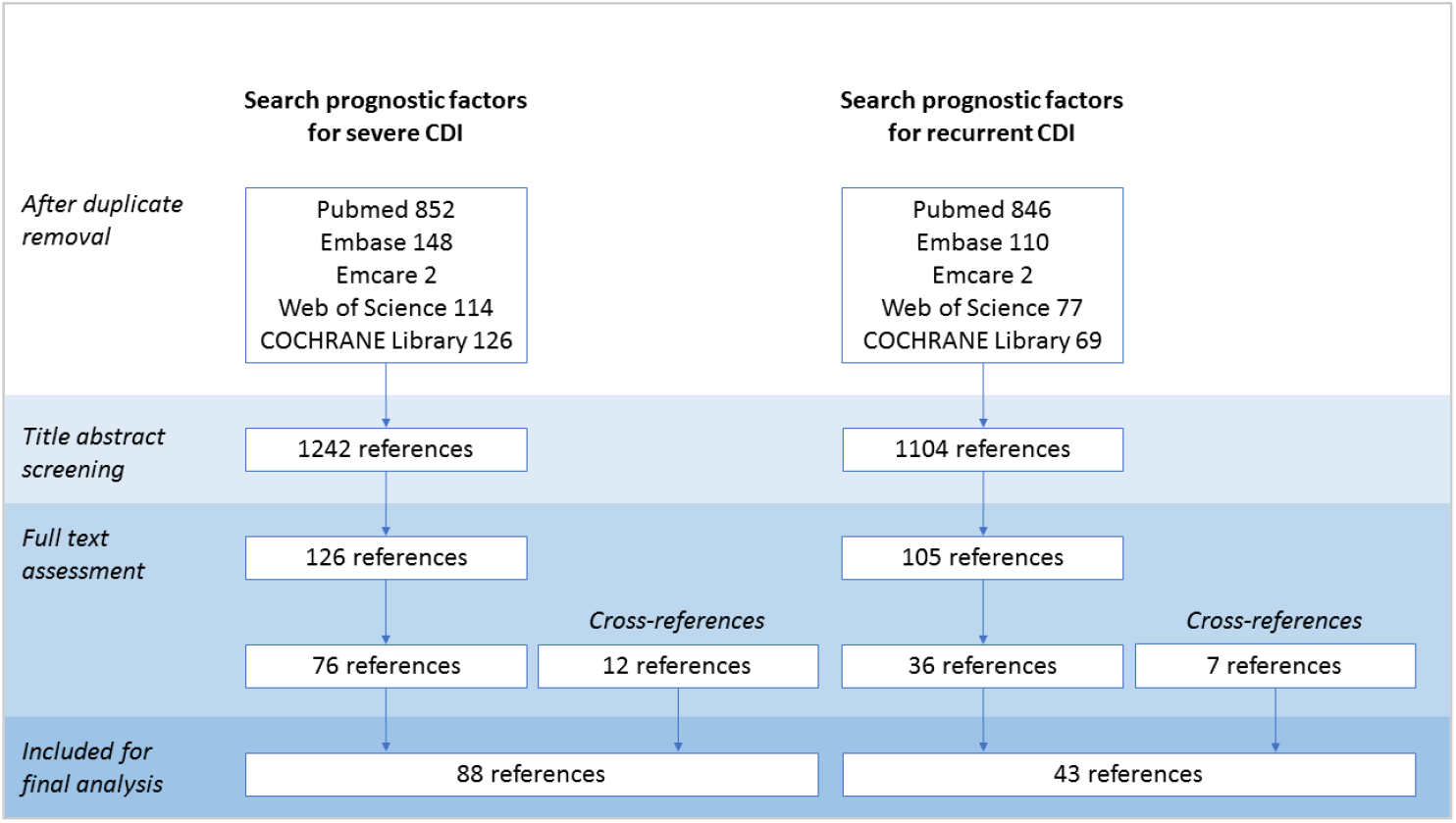
Flow diagram of the selection process of studies on prognostic factors for severe/recurrent CDI

### Prognostic factors for severe CDI

The overall quality of evidence ranged from very low to moderate, mainly due to the retrospective nature and small sample size of most studies. The prognostic factors studied in five or more articles are shown in **Table 1**.

**Table 1.**
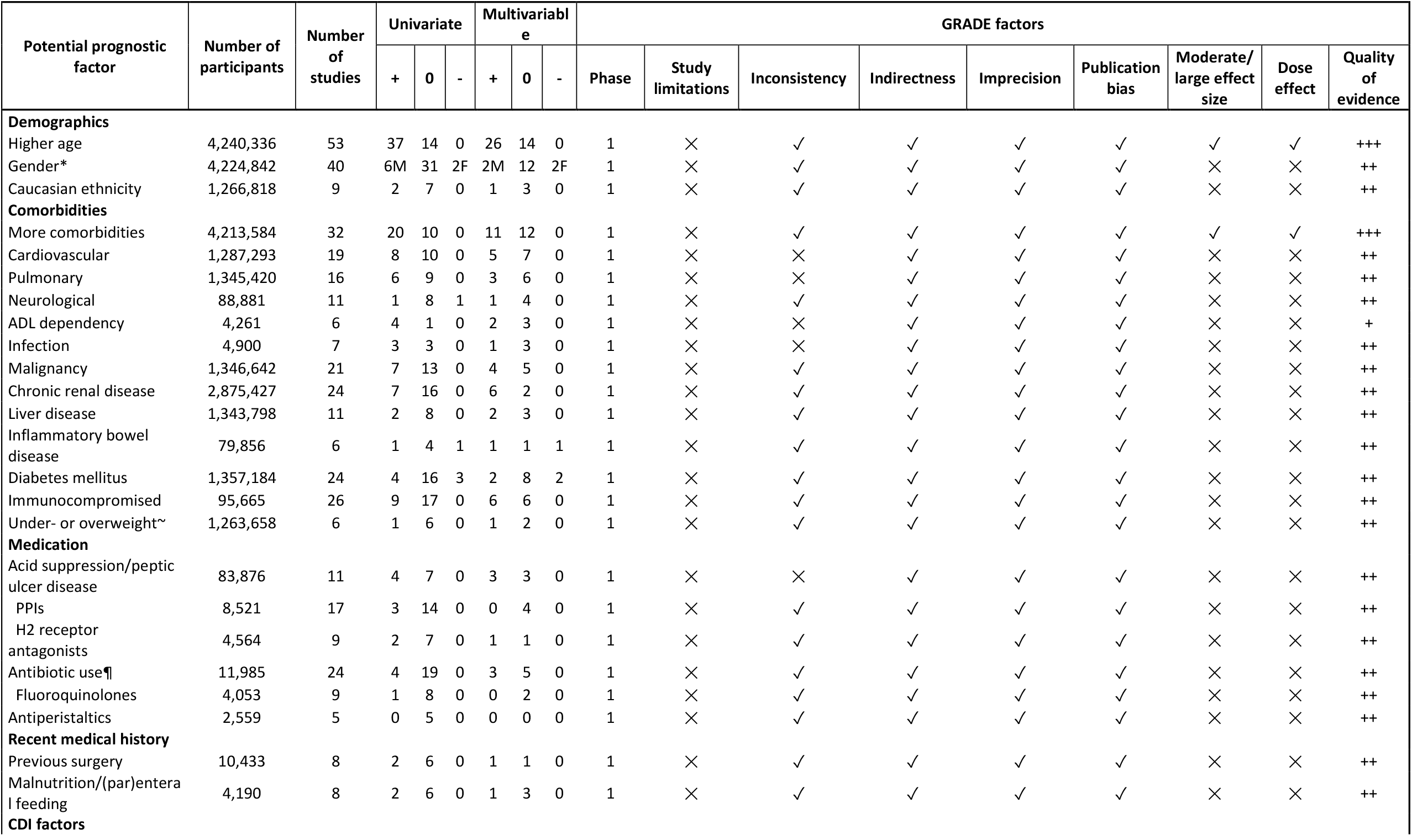
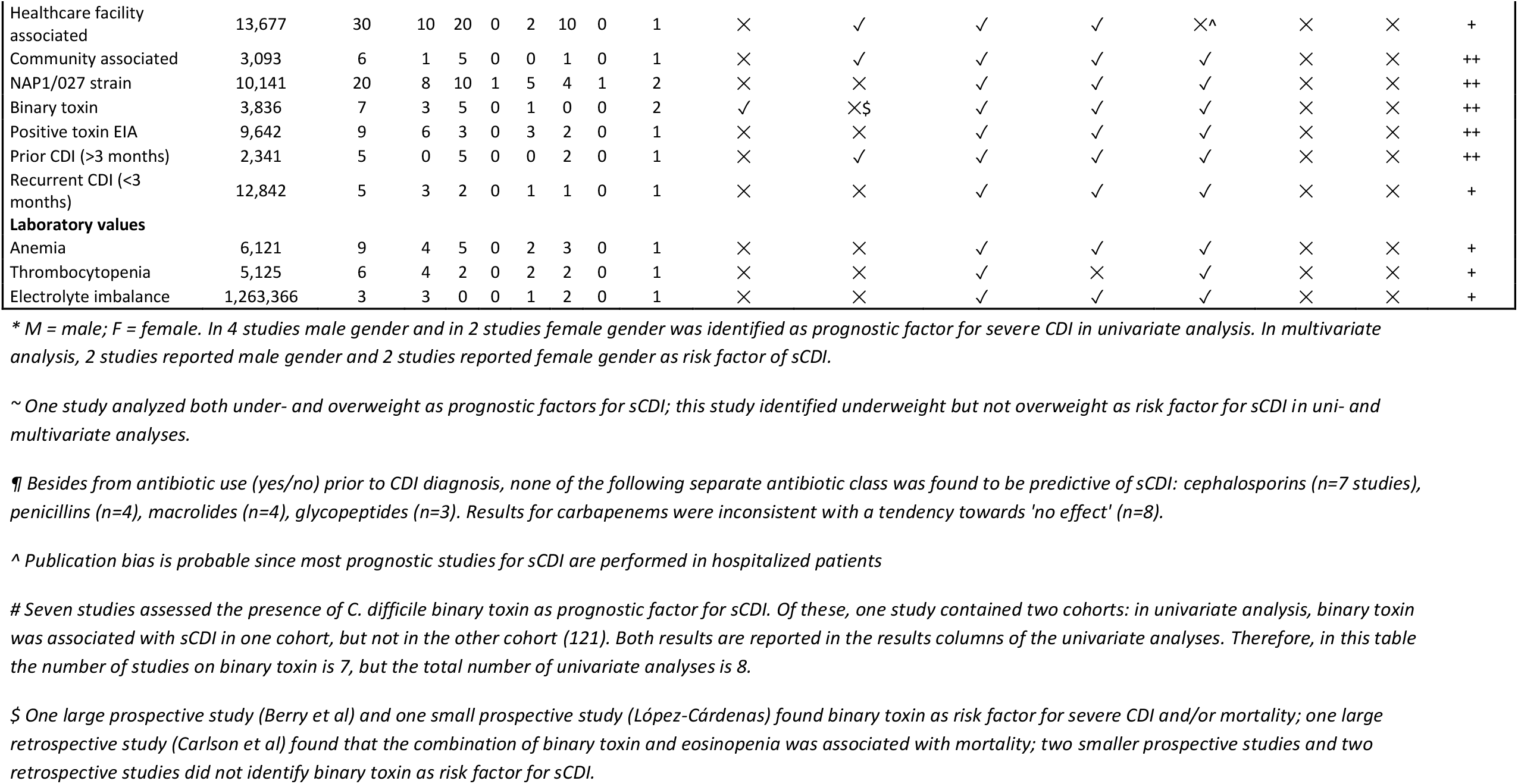
Potential prognostic factors for severe CDI. Phase, phase of investigation. For uni- and multivariate analyses: +, number of significant effects with a positive value; 0, number of non-significant effects; -, number of significant effects with a negative value. For GRADE factors: ✓, no serious limitations; ✗, serious limitations (or not present for moderate/large effect size, dose effect); unclear, unable to rate item based on available information. For overall quality of evidence: +, very low; ++, low; +++, moderate; ++++, high.

*Age* was the most studied prognostic factor, and was investigated in 53 of 88 included studies (20, 24, 25, 29, 38-42, 44, 46-49, 52, 54, 55, 57, 59, 61-64, 66, 70, 71, 75, 77, 78, 80, 81, 83, 85-88, 91-94, 96, 97, 100, 104, 106, 109-111, 113, 114, 116-118). Fifty-one studies performed a univariate analysis and 40 studies (also) a multivariable analysis; 37/51 studies reported *higher age* as risk factor for sCDI in univariate analysis, and 26/40 studies in multivariable analysis (overall moderate quality of evidence).

Thirty-two studies assessed the presence of *multiple comorbidities* as risk factor for sCDI (25, 38-42, 46-49, 52, 62, 66, 70, 72-74, 80, 81, 85, 87, 88, 91-94, 96, 97, 109, 114, 116, 117); 20/30 studies found an association between the presence of *multiple comorbidities* and sCDI in univariate analysis and 11/23 studies in multivariable analysis (moderate quality of evidence).

For both higher age and the presence of multiple comorbidities a dose effect was observed: the higher the age or the more concurrent disorders, the higher the risk of severe disease. No specific medical condition was associated with sCDI.

For clinical decision making, a specific cut-off value for age and number of comorbidities would be convenient. However, in many studies age was used as a continuous variable (31/53 studies) or varying cut-off values are used. The majority of studies found a higher risk for sCDI in patients older than 65-70 years. For comorbidity, different established or self-constructed comorbidity scores were reported, the Charlson Comorbidity Index (CCI) most frequently (161). Again, most studies used CCI as continuous variable. Of the studies that identified more comorbidities as risk factor for sCDI, only three used a cut-off value: one study reported a higher risk in patients with a CCI of ≥3, and two found a higher risk in patients with ≥2 comorbidities (88, 93, 96). Thus, no statement can be made on the association between exact numbers of comorbidities and severe CDI.

Twenty articles studied the association between the presence of NAP1/RT027 strain and sCDI (44-46, 56, 60, 61, 73, 74, 81, 84, 85, 88, 89, 91, 94, 96, 100, 101, 108, 111); 8/19 studies reported a higher risk of sCDI in patients with NAP1/027 strain in univariate analysis, and 5/10 in multivariate analysis. One study found that NAP1/027 was more prevalent in patients with mild disease compared to patients with sCDI (44).

Furthermore, seven studies assessed *C. difficile* binary toxin as risk factor for sCDI (45, 82, 91, 94, 117, 119, 121). Of these, three identified *C. difficile* binary toxin as a risk factor for sCDI in univariate analysis, and only 1/1 study in multivariate analysis. The overall quality of evidence of these studies was low. We scrutinized the separate studies on the presence of NAP1/RT027 strain and/or binary toxin by weighing the results by the quality of evidence per study, but this did not alter the conclusion.

Another interesting finding was that the majority of studies did not find an association between the use of *PPIs, H2 receptor antagonists* or *antibiotics* and the occurrence of sCDI in uni- and multivariate analyses. However, the quality of evidence was low.

In conclusion, only the factors *higher age (>65-70 years old)* and the presence of *multiple comorbidities* were consistently associated with sCDI (**Table 3**).

### Prognostic factors for recurrent CDI

The overall quality of evidence for the prognosis of recurrent CDI (rCDI) was low to moderate (**Table 2**). The majority of studies was retrospective, with a high to moderate risk of bias.

**Table 2.** Potential prognostic factors for recurrent CDI. Phase, phase of investigation. For uni- and multivariate analyses: +, number of significant effects with a positive value; 0, number of non-significant effects; -, number of significant effects with a negative value. For GRADE factors: ✓, no serious limitations; ✗, serious limitations (or not present for moderate/large effect size, dose effect); unclear, unable to rate item based on available information. For overall quality of evidence: +, very low; ++, low; +++, moderate; ++++, high

| Potential prognostic factor identified | Number of participants | Number of studies | Univariate |  |  | Multivariable |  |  | GRADE factors |  |  |  |  |  |  |  |  |
| --- | --- | --- | --- | --- | --- | --- | --- | --- | --- | --- | --- | --- | --- | --- | --- | --- | --- |
|  |  |  | + | 0 | - | + | 0 | - | Phase | Study limitations | Inconsistency | Indirectness | Imprecision | Publication bias | Moderate/<br>large effect size | Dose effect | Quality of evidence |
| Demographics |  |  |  |  |  |  |  |  |  |  |  |  |  |  |  |  |  |
| Higher age | 52,312 | 35 | 15 | 15 | 0 | 16 | 5 | 0 | 1 | × | × | ✓ | ✓ | ✓ | ✓ | ✓ | +++ |
| Gender | 7,074 | 8 | 0 | 7 | 0 | 0 | 1 | 0 | 1 | × | ✓ | ✓ | ✓ | ✓ | × | × | ++ |
| Male | 15,769 | 10 | 2 | 7 | 0 | 1 | 1 | 0 | 1 | × | ✓ | ✓ | ✓ | ✓ | × | × | ++ |
| Female | 5,833 | 12 | 2 | 9 | 0 | 1 | 2 | 0 | 1 | × | ✓ | ✓ | ✓ | ✓ | × | × | ++ |
| Community-acquired CDI | 13,837 | 7 | 1 | 5 | 1 | 1 | 1 | 0 | 1 | × | ✓ | ✓ | ✓ | ✓ | × | × | ++ |
| Hospital-acquired CDI | 10,468 | 10 | 5 | 5 | 0 | 2 | 2 | 0 | 1 | × | × | ✓ | ✓ | ✓ | × | × | ++ |
| Caucasian race | 13,335 | 3 | 2 | 1 | 0 | 1 | 0 | 0 | 1 | × | × | ✓ | ✓ | ✓ | × | × | + |
| CDI factors |  |  |  |  |  |  |  |  |  |  |  |  |  |  |  |  |  |
| Severe CDI | 8,796 | 10 | 2 | 7 | 0 | 2 | 2 | 0 | 1 | × | ✓ | ✓ | × | ✓ | × | × | + |
| Ribotype 027 strain | 6,921 | 5 | 2 | 3 | 0 | 1 | 2 | 0 | 1 | × | × | ✓ | ✓ | ✓ | × | × | ++ |
| Recurrent CDI episode (<3 months) | 20,868 | 8 | 2 | 3 | 0 | 3 | 1 | 0 | 1 | × | × | ✓ | ✓ | ✓ | × | × | ++ |
| Comorbidities |  |  |  |  |  |  |  |  |  |  |  |  |  |  |  |  |  |
| Charlson comorbidity index | 30,045 | 13 | 2 | 7 | 0 | 2 | 4 | 0 | 1 | × | ✓ | ✓ | ✓ | ✓ | × | × | ++ |
| Cardiovascular (CHF/MI) | 20,160 | 11 | 4 | 7 | 0 | 0 | 1 | 0 | 1 | × | ✓ | ✓ | × | ✓ | × | × | ++ |
| Peripheral vascular disease | 18,318 | 6 | 2 | 4 | 0 | 0 | 0 | 0 | 1 | × | × | ✓ | × | ✓ | × | × | + |
| Cerebrovascular (stroke) | 17,785 | 6 | 2 | 4 | 0 | 0 | 0 | 0 | 1 | × | ✓ | ✓ | ✓ | ✓ | × | × | ++ |
| (Chronic obstructive) Pulmonary disease | 19,908 | 11 | 1 | 10 | 0 | 0 | 0 | 0 | 1 | × | ✓ | ✓ | ✓ | ✓ | × | × | ++ |
| (Chronic) Renal disease/failure | 26,440 | 19 | 3 | 15 | 0 | 2 | 1 | 0 | 1 | × | ✓ | ✓ | ✓ | ✓ | × | × | +++ |
| Diabetes mellitus | 20,745 | 14 | 2 | 12 | 0 | 1 | 0 | 0 | 1 | × | ✓ | ✓ | ✓ | ✓ | × | × | +++ |
| Liver disease | 14,378 | 8 | 0 | 8 | 0 | 0 | 0 | 0 | 1 | × | ✓ | ✓ | ✓ | ✓ | × | × | ++ |
| Inflammatory bowel disease | 13,977 | 6 | 3 | 3 | 0 | 1 | 2 | 0 | 1 | × | × | ✓ | ✓ | ✓ | × | × | ++ |
| Dementia | 12,458 | 5 | 1 | 4 | 0 | 0 | 0 | 0 | 1 | × | × | ✓ | ✓ | ✓ | × | × | ++ |
| Malignancy | 13,685 | 10 | 0 | 10 | 0 | 0 | 0 | 0 | 1 | × | ✓ | ✓ | ✓ | ✓ | × | × | ++ |
| Solid malignancy | 10,078 | 4 | 1 | 3 | 0 | 0 | 0 | 0 | 1 | × | ✓ | ✓ | ✓ | ✓ | × | × | + |
| Hospital admission factors |  |  |  |  |  |  |  |  |  |  |  |  |  |  |  |  |  |
| Prior hospitalization (<3 months) | 31,952 | 8 | 5 | 2 | 0 | 2 | 2 | 0 | 1 | × | × | ✓ | ✓ | ✓ | × | × | ++ |
| Requirement of ICU admission | 28,356 | 8 | 2 | 4 | 0 | 0 | 3 | 0 | 1 | × | ✓ | ✓ | ✓ | ✓ | × | × | ++ |
| Increased length of stay (>10 days) | 2,464 | 5 | 3 | 2 | 0 | 1 | 2 | 0 | 1 | × | × | ✓ | ✓ | ✓ | × | × | + |
| Medication |  |  |  |  |  |  |  |  |  |  |  |  |  |  |  |  |  |
| Concomitant non-CDI AB | 7,980 | 8 | 2 | 4 | 0 | 2 | 3 | 0 | 1 | × | × | ✓ | ✓ | ✓ | × | × | + |
| Proton pump inhibitor | 43,653 | 23 | 6 | 14 | 1 | 8 | 3 | 1 | 1 | × | × | ✓ | ✓ | ✓ | × | × | ++ |
| PPI prior to CDI | 14,308 | 5 | 2 | 3 | 0 | 0 | 0 | 0 | 1 | × | × | ✓ | ✓ | ✓ | × | × | + |
| PPI during/after CDI | 11,785 | 7 | 6 | 1 | 0 | 2 | 2 | 0 | 1 | × | ✓ | ✓ | ✓ | ✓ | × | × | ++ |
| H2 receptor antagonist | 29,998 | 5 | 2 | 2 | 0 | 2 | 1 | 0 | 1 | × | ✓ | ✓ | ✓ | ✓ | × | × | ++ |
| Steroids / immunosuppression | 11,429 | 10 | 3 | 6 | 0 | 2 | 3 | 0 | 1 | × | ✓ | ✓ | ✓ | ✓ | × | × | ++ |
| Probiotics | 29,847 | 4 | 3 | 0 | 0 | 2 | 1 | 0 | 1 | × | ✓ | ✓ | ✓ | ✓ | × | × | ++ |
| Chemotherapy | 20,494 | 8 | 0 | 7 | 0 | 1 | 1 | 0 | 1 | × | ✓ | ✓ | ✓ | ✓ | × | × | ++ |
| Parenteral feeding | 2,988 | 7 | 1 | 6 | 0 | 0 | 0 | 0 | 1 | × | ✓ | ✓ | ✓ | × | × | × | + |
| **Inciting AB** |  |  |  |  |  |  |  |  |  |  |  |  |  |  |  |  |  |
| Aminopenicilin | 10,435 | 5 | 1 | 4 | 0 | 0 | 0 | 0 | 1 | × | ✓ | ✓ | ✓ | ✓ | × | × | ++ |
| Cephalosporin | 4,086 | 4 | 0 | 4 | 0 | 0 | 0 | 0 | 1 | × | ✓ | ✓ | ✓ | ✓ | × | × | ++ |
| Fluorquinolones | 15,657 | 12 | 3 | 9 | 0 | 2 | 0 | 0 | 1 | × | ✓ | ✓ | ✓ | ✓ | × | × | ++ |
| Aminoglycoside | 10,271 | 4 | 1 | 3 | 0 | 0 | 0 | 0 | 1 | × | ✓ | ✓ | ✓ | ✓ | × | × | ++ |
| Clindamycin | 10,679 | 5 | 2 | 3 | 0 | 1 | 0 | 0 | 1 | × | × | ✓ | ✓ | ✓ | × | × | + |
| Macrolide | 10,435 | 5 | 1 | 4 | 0 | 0 | 0 | 0 | 1 | × | ✓ | ✓ | ✓ | ✓ | × | × | ++ |
| Carbapenem/meropenem | 9,938 | 7 | 0 | 7 | 0 | 0 | 0 | 0 | 1 | × | ✓ | ✓ | ✓ | ✓ | × | × | ++ |
| **Laboratory values** |  |  |  |  |  |  |  |  |  |  |  |  |  |  |  |  |  |
| Leukocytes (mean/peak) | 22,954 | 17 | 6 | 10 | 0 | 1 | 3 | 0 | 1 | × | ✓ | ✓ | ✓ | ✓ | × | × | ++ |
| Albumin (mean/low) | 18,737 | 12 | 4 | 8 | 0 | 1 | 2 | 0 | 1 | × | ✓ | ✓ | ✓ | ✓ | × | × | ++ |
| Creatinine (mean/peak) | 19,651 | 14 | 1 | 12 | 0 | 1 | 2 | 0 | 1 | × | ✓ | ✓ | ✓ | ✓ | × | × | +++ |
| C-reactive protein | 9,805 | 5 | 2 | 2 | 0 | 0 | 0 | 0 | 1 | × | × | ✓ | ✓ | ✓ | × | × | + |
| **Symptoms** |  |  |  |  |  |  |  |  |  |  |  |  |  |  |  |  |  |
| Fever | 3,740 | 9 | 3 | 6 | 0 | 2 | 0 | 0 | 1 | × | ✓ | ✓ | ✓ | ✓ | × | × | ++ |
| Abdominal pain | 5,756 | 5 | 2 | 3 | 0 | 0 | 0 | 0 | 1 | × | × | ✓ | ✓ | ✓ | × | × | + |

**Table 3.** Summary of findings

| Prognostic factor | Quality of evidence |
| --- | --- |
| *Severe CDI* |  |
| Higher age (>65-70 years old) | moderate |
| Presence of multiple comorbidities | moderate |
| *Recurrent CDI* |  |
| Higher age (>65-70 years old) | moderate |
| Previous recurrence of CDI (<3 months) | moderate |
| Healthcare-associated CDI | low |
| Prior hospitalization (<3 months) | low |
| Proton pump inhibitors started during/after CDI diagnosis | low |

*Higher age (≥65 years old)* is the most studied factor and was investigated in 35/43 included studies (9, 10, 12-15, 109, 111, 122, 123, 125, 126, 128, 130-134, 136-141, 143-153), 15/30 studies identified higher age as risk factor for rCDI in univariate analysis, and 16/31 studies in multivariable analysis. Moreover, higher age was the only factor for which we found a moderate to large effect size and a dose-dependent effect.

Eight studies assessed whether a *previous CDI recurrence in the preceding 3 months* was a risk factor for a subsequent rCDI, and showed inconsistent results (9, 10, 15, 111, 125, 133, 140, 145). Two prospective studies of higher quality found a clear association of previous recurrence with the risk of a subsequent rCDI (111, 140). Also, data of the pivotal trials on fidaxomicin and bezlotoxumab indicate that recurrence rates are higher in patients with any previous CDI episode or patients that fulfill criteria for rCDI (156-160). Since these trials are considered high quality studies, we have upgraded the level of evidence from low to moderate (Table 3). Finally, we consider two or more episodes of CDI as risk factor for a subsequent rCDI.

*Healthcare-acquired CDI* was associated with rCDI in univariate analyses of 5/10 studies, and in multivariable analyses of 2/4 studies (9, 14, 111, 125, 128, 141, 144, 145, 149, 150). This is also reflected by the correlation of *prior hospitalization (<3 months)* with a recurrent episode of CDI (13, 15, 125, 128, 133, 138, 143, 149). The two largest cohort studies showed a significant association between prior hospitalization and rCDI in uni- and multivariable analysis (128, 149). Of note, one large cohort study reported a protective effect of community-acquired CDI for the development of rCDI (128).

*Proton pump inhibitor use* was studied in 23 included studies (9, 13, 14, 109, 122, 123, 130-135, 137, 139, 141, 145, 147-151, 153, 155). The results show no clear association between PPIs and rCDI in univariate analyses, while in multivariable analyses there appears to be an association. Of note, some studies made a distinction between the moment PPI was started, i.e. prior to initial CDI episode or during/shortly after the initial episode (12, 13, 15, 123, 125, 146, 149, 155); PPI prescribed during or shortly after initial CDI appears to be associated with an increased risk of rCDI (effect found in 6/7 studies in univariate analysis, and in 2/4 studies in multivariable analysis; low quality of evidence).

Eight studies reported on *non-CDI antibiotic use* after initial CDI diagnosis as a risk factor, but results were inconsistent (12, 123, 129, 136, 141, 145, 146, 148); 4/6 studies found an association between non-CDI antibiotic use and rCDI in univariate analysis, and 2/5 studies in multivariable analysis. A high-quality prospective study showed no significant association between antibiotic use and rCDI in both uni- and multivariate analysis (136). Several other studies investigated whether specific antibiotics that incited the primary CDI episode, were associated with recurrence; none of these antibiotic drugs was associated with rCDI. In conclusion, nor non-CDI antibiotic use prior to the primary CDI, nor concomitant non-CDI antibiotics used during the primary CDI episode, appeared to be convincing predictors for rCDI.

Although various definitions of sCDI are used in prognostic studies, most studies did not find severity of CDI to be a prognostic factor for rCDI (9, 123, 126, 131, 134, 140-142, 147, 150); 2/7 studies identified sCDI as risk factor for rCDI in univariate analysis, and 2/4 in multivariable analysis. In the MODIFY trials, sCDI was a prespecified risk factor for recurrence, but in the placebo arm the recurrence rate was lower in the sCDI subgroup (22.4%) than in the non-CDI subgroup (27.5%) (158). Since we expected sCDI to be a risk factor for rCDI, re-assessment of included studies was performed by a third researcher (CV-G); this did not alter our results. In conclusion, we found insufficient evidence to consider sCDI a risk factor for rCDI.

Several studies investigated specific comorbidities, including chronic renal failure, diabetes mellitus, and cardiovascular disease as risk factors for rCDI (9, 12-15, 111, 122, 123, 125, 126, 128, 130-134, 136, 138-154). Our review does not show any of these comorbidities to be clearly associated with an increased risk for rCDI. Even when combined into a robust score such as the Charlson Comorbidity Index, we did not identify a clear association (13, 65, 122, 130, 132-134, 140, 145, 146, 148, 150, 152). Since results of the uni- and multivariable analyses of studies reporting in renal failure were conflicting, these studies were re-assessed by a third researcher. This led to identical findings.

Immunocompromised status was a prespecified risk factor in the MODIFY trials; though in the placebo arm the recurrence rate in immunocompromised patients did not differ from that in immunocompetent patients (27.5% vs 26.6%) (158). A post-hoc analysis of the pivotal fidaxomicin trials investigated fever, leukocytosis and renal failure as prognostic markers, and identified renal failure as the only significant predictor for recurrence (RR, 1.45; 95% CI, 1.05–2.02). Overall, we found insufficient evidence to consider other comorbidities a risk factor for rCDI.

The ribotype 027 strain was not clearly associated with recurrence. However, only five studies reported on this ribotype as a possible risk factor (111, 123, 129, 145, 148); 2/5 studies found that the presence of a RT027 strain was associated with sCDI in univariate analysis, and only 1/3 studies in multivariable analysis. Since these results were inconsistent when compared to a previous systematic review, results were re-assessed by a third researcher (162). This reassessment did not result in different conclusions.

Laboratory parameters studied include white blood cell counts, and levels albumin, creatinine and C-reactive protein. None of these parameters was clearly associated with rCDI (10, 12-15, 111, 134, 141, 144-147, 149).

Our findings show the use of probiotics as prophylaxis for CDI seemed to be associated with rCDI (13, 14, 123, 133). However, this association is based on a small number of studies (n=4) and might be confounded by the fact that probiotics are started in patients with a high risk of rCDI and/or concomitant antibiotic use.

## Conclusions

In this systematic review, we applied the GRADE methodology to weigh the evidence on prognostic factors for sCDI and rCDI (37). We found that the most important risk factors for sCDI were higher age (≥65-70 years old) and presence of multiple comorbidities. Higher age was also a prognostic factor for rCDI. Other factors associated with an increased risk of rCDI were the acquisition of CDI as a healthcare-associated infectionand prior hospitalization and the start of PPIs during/after CDI diagnosis. The evidence for having a previous recurrence of CDI as risk factor for rCDI was less strong.

Our systematic review on prognostic factors resulted in several expected and unexpected findings when compared with previous reviews rCDI (162-164). Our findings are in line with two recently published systematic reviews on predictors of sCDI that reported age and presence of comorbidities as the two most important severity predictors for CDI (162, 165). Similarly, previous reviews on prognostic factors for rCDI found that older age, prior CDI and PPI therapy were risk factors for rCDI (162-165). However, PPI therapy in general did not show a clear association in our study.

In contrast, we could not clearly identify several previously reported risk factors for sCDI or rCDI, such as infection caused by a ribotype 027/NAP1 strain, antibiotic use, PPI use prior to CDI diagnosis, and the presence of chronic or end-stage renal disease (162, 163). One possible explanation might be that many reviews excluded studies reporting solely univariate analyses. This might lead to an overestimation of the effect of certain prognostic factors, since many studies only include variables with a significant association in univariate analysis in their multivariable analysis. Another explanation for our findings might be index event bias: this bias arises in studies that select patients based on the occurrence of an index event, and can lead to ‘negative’ or even paradoxical findings in recurrence risk research (166). Here, we found that chronic renal failure did not appear to be associated with rCDI. Previous research shows that patients with kidney failure are prone to developing a primary CDI, but the direct association between kidney disease and rCDI is less clear (163, 167). Since reviews on rCDI select studies based on the occurrence of a primary CDI, this might explain why renal disease is not a strong predictor of rCDI in previous reviews, and not a significant risk factor in the current review.

Finally, when studying IBD as prognostic factor for sCDI/rCDI, another difficulty in interpretation arises, since symptoms of IBD are similar to those of CDI. Furthermore, patients with IBD are at higher risk of CDI and infection and colonization with *C. difficile* can induce an exacerbation of IBD (168). Also, diagnostic tests do not always allow to differentiate between *C. difficile* infection or colonization. Many clinicians pragmatically start antibiotic treatment in IBD patients without endoscopy. Subsequent false-positive CDI diagnoses may contribute to an overestimation of CDI (recurrence) risk. Aforementioned factors may have contributed to differences between the results of our current review and previous publications.

Our review has several strengths. First, we used a broad search strategy and included a large number of studies, minimizing the risk of missing data. Second, articles were selected based on strict predefined criteria by two independent researchers (TvR and RO). Third, the quality of the studies was judged by the structured prognostic GRADE approach. To our knowledge this approach has not been previously used for prognostic factors for sCDI and rCDI. Also, we reported results of both uni- and multivariable analyses. When results of the uni- and multivariable analyses of different studies were conflicting, we acknowledged this disparity and results were re-assessed by a third researcher. Reassessment was performed on the studies reporting the presence of RT027 strain, sCDI and renal disease as possible risk factors for rCDI, but this did not result in different conclusions. Finally, some disparity can be explained by the large number of studies where non-significant univariate prognostic factors were not included in the multivariate models.

A limitation is the generally low quality of available studies. Most studies have not performed a sample size calculation, allowing for possible type I and type II statistical errors. As many studies were retrospective, it was not always clear whether certain factors were already present before the occurrence of sCDI/rCDI (i.e. truly prognostic factors) or whether they co-existed. A second important limitation is the subjectivity how to weight different quality studies, which also resulted in stimulating discussions in our working group.

Identifying prognostic factors for sCDI and rCDI could aid clinicians to make an optimal treatment decision to reduce the risk of recurrent disease and decrease CDI complications. The findings of this review are especially relevant for rCDI: a risk stratification strategy may allow for selective use of more expensive novel treatments with less risk of recurrence. To implement risk stratification of in clinical practice, simple risk tools are needed. Currently, several prediction models or risk scores for scDI and rCDI have been developed (9-30). However, none has gained widespread clinical implementation. Based on the low quality of the studies and small effects of identified prognostic factors, it is not surprising that validation of prediction tools in external cohorts show disappointing results (169-171). Additional explanations are the heterogeneity and multimorbidity of the CDI population and variation in study setting, diagnostic criteria and CDI treatment regiments. The current review may be used as starting point for the development of a risk tool with a better performance in the overall CDI population. We suggest that future clinical trials on CDI treatment use standardized definitions for recurrence and severity and systematically collect and report the risk factors that we have discussed in this review, to allow for meaningful meta-analysis on risk factors using data of high-quality trials.

In conclusion, our approach identified higher age and the presence of multiple comorbidities as prognostic factors for sCDI. The only identified risk factors for rCDI were higher age, healthcare-associated CDI, prior hospitalization (<3 months), PPIs started during/after CDI diagnosis and rCDI. Some variables that were found as risk factors for sCDI/rCDI in previous reviews, were not confirmed in the current review, which can be attributed to differences in methodology. Risk stratification for sCDI/rCDI may contribute to a more personalized and optimal treatment for patients with CDI.

## Data Availability

Not applicable (systematic review)

## Transparency declaration

### Contribution

JP and EK conceived the study on behalf of the *ESCMID Study Group for Clostridioides difficile* (ESGCD). JP designed the search strategy. TvR and RO selected the studies and acquired the data. TvR, RO, CV-G and JK graded selected studies. TvR, RO and OD analyzed the data. TvR and RO drafted the manuscript. All authors critically revised the manuscript and approved the final version.

## Supplementary data

### PICO’s

Two PICO’s were formulated:

Prognosis severe CDI:

P: patients with CDI

I: presence of clinical and laboratory parameters

C: absence of clinical and laboratory parameters

O: severe CDI

Prognosis recurrent CDI:

P: patients with CDI

I: presence of clinical and laboratory parameters

C: absence of clinical and laboratory parameters

O: recurrent CDI

### Literature search prognosis severe CDI - Pubmed

((“Clostridium difficile Infection” [tw] OR “Clostridium difficile Infections” [tw] OR “C difficile Infection” [tw] OR “C difficile Infections” [tw] OR “clostridioides difficile infection” [tw] OR ((“Clostridium difficile” [majr] OR “Clostridium difficile” [ti] OR “C difficile” [ti] OR “c diff” [ti] OR “clostridioides difficile” [ti]) AND (“Clostridium Infections” [majr] OR “Clostridium Infection” [ti] OR “Clostridium Infection” [ti])) OR ((“Clostridium difficile” [majr] OR “Clostridium difficile” [ti] OR “C difficile” [ti] OR “difficile” [ti]) AND (“Clostridium Infections” [majr] OR “Clostridium Infection” [ti] OR “Clostridium Infection” [ti] OR “Infection” [majr] OR “infection” [ti] OR “infections” [ti] OR infect*[ti] OR “inflammation” [ti] OR inflammat*[ti] OR “Enterocolitis” [majr] OR “Enterocolitis” [ti] OR “colitis” [ti] OR “Digestive System Diseases” [majr])))

**AND**

(“Prognosis” [Mesh] OR prognos*[tw] OR “prognostic factor” [tw] OR “prognostic factors” [tw] OR “clinical parameter” [tw] OR “clinical parameters” [tw] OR “laboratory parameter” [tw] OR “laboratory parameters” [tw] OR “Age Factors” [mesh] OR “Age Factor” [tw] OR “Age Factors” [tw] OR “Age” [tiab] OR “Age Reporting” [tw] OR “Leukocytosis” [Mesh] OR “Leukocytosis” [tw] OR “Pleocytosis” [tw] OR “leucocytosis” [tw] OR “Leukemoid Reaction” [tw] OR “Lymphocytosis” [tw] OR (decreas*[tw] AND (“Serum Albumin” [mesh] OR “blood albumin” [tw] OR “serum albumin” [tw])) OR ((“rise” [tw] OR “rising” [tw] OR rise*[tw] OR increas*[tw]) AND (“serum creatinine” [tw] OR “Creatinine/blood” [mesh])) OR “severe underlying disease” [tw] OR “severe underlying diseases” [tw] OR “severe underlying illness” [tw] OR “severe underlying illnesses” [tw] OR “severe underlying condition” [tw] OR “severe underlying conditions” [tw] OR “Immunologic Deficiency Syndromes” [Mesh] OR “Immunologic Deficiency” [tw] OR “Immune Deficiency” [tw] OR “immunodeficiency” [tw] OR “Immunologic Deficiencies” [tw] OR “Immune Deficiencies” [tw] OR “immunodeficiencies” [tw] OR “Immunocompromised Host” [Mesh] OR “Immunocompromised” [tw] OR (strain*[tw] AND ribotype*[tw]))

**AND**

(“severe clostridium difficile” [tw] OR “severe cdi” [tw] OR “severe cd” [tw] OR “disease severity” [tw] OR “illness severity” [tw] OR “severity of disease” [tw] OR “severity of illness” [tw] OR “severe” [tw] OR “severity” [tw] OR sever*[tw])

**NOT**

((“Case Reports” [ptyp] OR “case report” [ti]) NOT (“Review” [ptyp] OR “review” [ti] OR “Clinical Study” [ptyp] OR “trial” [ti] OR “RCT” [ti])) NOT (“Animals” [mesh] NOT “Humans” [mesh])

**AND**

english[la])

### Literature search prognosis recurrent CDI - Pubmed

((“Clostridium difficile Infection” [tw] OR “Clostridium difficile Infections” [tw] OR “C difficile Infection” [tw] OR “C difficile Infections” [tw] OR “clostridioides difficile infection” [tw] OR ((“Clostridium difficile” [majr] OR “Clostridium difficile” [ti] OR “C difficile” [ti] OR “c diff” [ti] OR “clostridioides difficile” [ti]) AND (“Clostridium Infections” [majr] OR “Clostridium Infection” [ti] OR “Clostridium Infection” [ti])) OR ((“Clostridium difficile” [majr] OR “Clostridium difficile” [ti] OR “C difficile” [ti] OR “difficile” [ti]) AND (“Clostridium Infections” [majr] OR “Clostridium Infection” [ti] OR “Clostridium Infection” [ti] OR “Infection” [majr] OR “infection” [ti] OR “infections” [ti] OR infect*[ti] OR “inflammation” [ti] OR inflammat*[ti] OR “Enterocolitis” [majr] OR “Enterocolitis” [ti] OR “colitis” [ti] OR “Digestive System Diseases” [majr])))

**AND**

(“Prognosis” [Mesh] OR prognos*[tw] OR “prognostic factor” [tw] OR “prognostic factors” [tw] OR “clinical parameter” [tw] OR “clinical parameters” [tw] OR “laboratory parameter” [tw] OR “laboratory parameters” [tw] OR “Age Factors” [mesh] OR “Age Factor” [tw] OR “Age Factors” [tw] OR “Age” [tiab] OR “Age Reporting” [tw] OR “severe underlying disease” [tw] OR “severe underlying diseases” [tw] OR “severe underlying illness” [tw] OR “severe underlying illnesses” [tw] OR “severe underlying condition” [tw] OR “severe underlying conditions” [tw] OR “Immunologic Deficiency Syndromes” [Mesh] OR “Immunologic Deficiency” [tw] OR “Immune Deficiency” [tw] OR “immunodeficiency” [tw] OR “Immunologic Deficiencies” [tw] OR “Immune Deficiencies” [tw] OR “immunodeficiencies” [tw] OR “Immunocompromised Host” [Mesh] OR “Immunocompromised” [tw] OR (strain*[tw] AND ribotype*[tw]) OR ((continued use*[tw] OR continuing use*[tw] OR prolonged use*[tw]) AND (antibiotic*[tw] OR antibacterial*[tw] OR anti-biotic*[tw] OR anti-bacterial*[tw])) OR “renal failure” [tw] OR “kidney failure” [tw] OR “Renal Insufficiency” [Mesh] OR “Renal Insufficiency” [tw] OR “Kidney Insufficiency” [tw] OR “kidney injury” [tw] OR “renal injury” [tw] OR previous infection*[tw] OR “Proton Pump Inhibitors” [Mesh] OR “Proton Pump Inhibitors” [Pharmacological Action] OR “Proton Pump Inhibitors” [tw] OR “Proton Pump Inhibitor” [tw] OR “initial severity” [tw] OR “initial disease severity” [tw] OR “initial illness severity” [tw] OR (strain*[tw] AND ribotype*[tw]))

**AND**

(“recurrent clostridium difficile” [tw] OR “recurrent cdi” [tw] OR “recurrent cd” [tw] OR “Recurrence” [Mesh] OR recurr*[tw])

**NOT**

((“Case Reports” [ptyp] OR “case report” [ti]) NOT (“Review” [ptyp] OR “review” [ti] OR “Clinical Study” [ptyp] OR “trial” [ti] OR “RCT” [ti])) NOT (“Animals” [mesh] NOT “Humans” [mesh])

**AND**

english[la])

## References

1. Lessa FC, Mu Y, Bamberg WM, Beldavs ZG, Dumyati GK, Dunn JR, et al. Burden of Clostridium difficile infection in the United States. N Engl J Med. 2015;372(9):825–34.

2. McDonald LC, Gerding DN, Johnson S, Bakken JS, Carroll KC, Coffin SE, et al. Clinical Practice Guidelines for Clostridium difficile Infection in Adults and Children: 2017 Update by the Infectious Diseases Society of America (IDSA) and Society for Healthcare Epidemiology of America (SHEA). Clin Infect Dis. 2018;66(7):987–94.

3. Debast SB, Bauer MP, Kuijper EJ, European Society of Clinical M, Infectious D. European Society of Clinical Microbiology and Infectious Diseases: update of the treatment guidance document for Clostridium difficile infection. Clin Microbiol Infect. 2014;20 Suppl 2:1–26.

4. Ooijevaar RE, van Beurden YH, Terveer EM, Goorhuis A, Bauer MP, Keller JJ, et al. Update of treatment algorithms for Clostridium difficile infection. Clinical microbiology and infection: the official publication of the European Society of Clinical Microbiology and Infectious Diseases. 2018;24(5):452–62.

5. Beinortas T, Burr NE, Wilcox MH, Subramanian V. Comparative efficacy of treatments for Clostridium difficile infection: a systematic review and network meta-analysis. Lancet Infect Dis. 2018;18(9):1035–44.

6. Wilcox MH, Gerding DN, Poxton IR, Kelly C, Nathan R, Birch T, et al. Bezlotoxumab for Prevention of Recurrent Clostridium difficile Infection. N Engl J Med. 2017;376(4):305–17.

7. Ianiro G, Masucci L, Quaranta G, Simonelli C, Rizzatti G, Lopetuso L, et al. Randomized Clinical Trial: Single-Infusion Fmt Versus Multiple-Infusion Fmt for the Treatment of Severe C. Difficile Infection. Digest Liver Dis. 2018;50(2):E66–E7.

8. van Nood E, Vrieze A, Nieuwdorp M, Fuentes S, Zoetendal EG, de Vos WM, et al. Duodenal infusion of donor feces for recurrent Clostridium difficile. N Engl J Med. 2013;368(5):407–15.

9. Cobo J, Merino E, Martinez C, Cozar-Llisto A, Shaw E, Marrodan T, et al. Prediction of recurrent clostridium difficile infection at the bedside: the GEIH-CDI score. Int J Antimicrob Agents. 2018;51(3):393–8.

10. D’Agostino RB, Sr., Collins SH, Pencina KM, Kean Y, Gorbach S. Risk estimation for recurrent Clostridium difficile infection based on clinical factors. Clin Infect Dis. 2014;58(10):1386–93.

11. Zilberberg MD, Reske K, Olsen M, Yan Y, Dubberke ER. Development and validation of a recurrent Clostridium difficile risk-prediction model. J Hosp Med. 2014;9(7):418–23.

12. Larrainzar-Coghen T, Rodriguez-Pardo D, Puig-Asensio M, Rodriguez V, Ferrer C, Bartolome R, et al. First recurrence of Clostridium difficile infection: clinical relevance, risk factors, and prognosis. European journal of clinical microbiology & infectious diseases: official publication of the European Society of Clinical Microbiology. 2016;35(3):371–8.

13. Reveles KR, Mortensen EM, Koeller JM, Lawson KA, Pugh MJV, Rumbellow SA, et al. Derivation and Validation of a Clostridium difficile Infection Recurrence Prediction Rule in a National Cohort of Veterans. Pharmacotherapy. 2018;38(3):349–56.

14. Viswesh V, Hincapie AL, Yu M, Khatchatourian L, Nowak MA. Development of a bedside scoring system for predicting a first recurrence of Clostridium difficile-associated diarrhea. Am J Health Syst Pharm. 2017;74(7):474–82.

15. Hebert C, Du H, Peterson LR, Robicsek A. Electronic health record-based detection of risk factors for Clostridium difficile infection relapse. Infect Control Hosp Epidemiol. 2013;34(4):407–14.

16. LaBarbera FD, Nikiforov I, Parvathenani A, Pramil V, Gorrepati S. A prediction model for Clostridium difficile recurrence. J Community Hosp Intern Med Perspect. 2015;5(1):26033.

17. Hu MY, Katchar K, Kyne L, Maroo S, Tummala S, Dreisbach V, et al. Prospective derivation and validation of a clinical prediction rule for recurrent Clostridium difficile infection. Gastroenterology. 2009;136(4):1206–14.

18. Abou Chakra CN, Pepin J, Valiquette L. Prediction tools for unfavourable outcomes in Clostridium difficile infection: a systematic review. PloS one. 2012;7(1):e30258.

19. Arora V, Kachroo S, Ghantoji SS, Dupont HL, Garey KW. High Horn’s index score predicts poor outcomes in patients with Clostridium difficile infection. The Journal of hospital infection. 2011;79(1):23–6.

20. Bhangu S, Bhangu A, Nightingale P, Michael A. Mortality and risk stratification in patients with Clostridium difficile-associated diarrhoea. Colorectal disease: the official journal of the Association of Coloproctology of Great Britain and Ireland. 2010;12(3):241–6.

21. Butt E, Foster JA, Keedwell E, Bell JE, Titball RW, Bhangu A, et al. Derivation and validation of a simple, accurate and robust prediction rule for risk of mortality in patients with Clostridium difficile infection. BMC Infect Dis. 2013;13:316.

22. Drew RJ, Boyle B. RUWA scoring system: a novel predictive tool for the identification of patients at high risk for complications from Clostridium difficile infection. The Journal of hospital infection. 2009;71(1):93–4; author reply 4-5.

23. Hensgens MP, Dekkers OM, Goorhuis A, LeCessie S, Kuijper EJ. Predicting a complicated course of Clostridium difficile infection at the bedside. Clin Microbiol Infect. 2014;20(5):O301–8.

24. Lungulescu OA, Cao W, Gatskevich E, Tlhabano L, Stratidis JG. CSI: a severity index for Clostridium difficile infection at the time of admission. The Journal of hospital infection. 2011;79(2):151–4.

25. Na X, Martin AJ, Sethi S, Kyne L, Garey KW, Flores SW, et al. A Multi-Center Prospective Derivation and Validation of a Clinical Prediction Tool for Severe Clostridium difficile Infection. PloS one. 2015;10(4):e0123405.

26. Rubin MS, Bodenstein LE, Kent KC. Severe Clostridium difficile colitis. Diseases of the colon and rectum. 1995;38(4):350–4.

27. Valiquette L, Pépin J, Do XV, Nault V, Beaulieu AA, Bédard J, et al. Prediction of complicated Clostridium difficile infection by pleural effusion and increased wall thickness on computed tomography. Clin Infect Dis. 2009;49(4):554–60.

28. Velazquez-Gomez I, Rocha-Rodriguez R, Toro DH, Gutierrez-Nuñez JJ, Gonzalez G, Saavedra S. A Severity Score Index for Clostridium difficile Infection. Infectious Diseases in Clinical Practice. 2008;16(6):376–8.

29. Welfare MR, Lalayiannis LC, Martin KE, Corbett S, Marshall B, Sarma JB. Co-morbidities as predictors of mortality in Clostridium difficile infection and derivation of the ARC predictive score. The Journal of hospital infection. 2011;79(4):359–63.

30. Zilberberg MD, Shorr AF, Micek ST, Doherty JA, Kollef MH. Clostridium difficile-associated disease and mortality among the elderly critically ill. Critical care medicine. 2009;37(9):2583–9.

31. Bauer MP, Kuijper EJ, van Dissel JT, European Society of Clinical M, Infectious D. European Society of Clinical Microbiology and Infectious Diseases (ESCMID): treatment guidance document for Clostridium difficile infection (CDI). Clin Microbiol Infect. 2009;15(12):1067–79.

32. J. van Prehn ER, E.H. Vogelzang, E. Bouza, A. Hristea, B. Guery, M. Krutova, T. Norén, F. Allerberger, J. Coia, A. Goorhuis, T. M. van Rossen, R.E. Ooijevaar, K. Burns, B.R. Scharvik Olesen, S. Tschudin-Sutter, M. H. Wilcox, M. Vehreschild, F. Fitzpatrick, E.J. Kuijper. European Society of Clinical Microbiology and Infectious Diseases: 2021 update on the treatment guidance document for Clostridioides difficile infection in adults. submitted to the ESCMID for (public) approval. 2021.

33. McDonald LC, Gerding DN, Johnson S, Bakken JS, Carroll KC, Coffin SE, et al. Clinical Practice Guidelines for Clostridium difficile Infection in Adults and Children: 2017 Update by the Infectious Diseases Society of America (IDSA) and Society for Healthcare Epidemiology of America (SHEA). Clinical infectious diseases: an official publication of the Infectious Diseases Society of America. 2018;66(7):e1–e48.

34. Surawicz CM, Brandt LJ, Binion DG, Ananthakrishnan AN, Curry SR, Gilligan PH, et al. Guidelines for diagnosis, treatment, and prevention of Clostridium difficile infections. Am J Gastroenterol. 2013;108(4):478–98; quiz 99.

35. Kuijper EJ, Coignard B, Tull P, difficile ESGfC, States EUM, European Centre for Disease P, et al. Emergence of Clostridium difficile-associated disease in North America and Europe. Clin Microbiol Infect. 2006;12 Suppl 6:2–18.

36. Hayden JA, van der Windt DA, Cartwright JL, Cote P, Bombardier C. Assessing bias in studies of prognostic factors. Ann Intern Med. 2013;158(4):280–6.

37. Huguet A, Hayden JA, Stinson J, McGrath PJ, Chambers CT, Tougas ME, et al. Judging the quality of evidence in reviews of prognostic factor research: adapting the GRADE framework. Syst Rev. 2013;2:71.

38. Alicino C, Giacobbe DR, Durando P, Bellina D, AM DIB, Paganino C, et al. Increasing incidence of Clostridium difficile infections: results from a 5-year retrospective study in a large teaching hospital in the Italian region with the oldest population. Epidemiology and infection. 2016;144(12):2517–26.

39. Ananthakrishnan AN, Guzman-Perez R, Gainer V, Cai T, Churchill S, Kohane I, et al. Predictors of severe outcomes associated with Clostridium difficile infection in patients with inflammatory bowel disease. Alimentary pharmacology & therapeutics. 2012;35(7):789–95.

40. Andrews CN, Raboud J, Kassen BO, Enns R. Clostridium difficile-associated diarrhea: predictors of severity in patients presenting to the emergency department. Canadian journal of gastroenterology = Journal canadien de gastroenterologie. 2003;17(6):369–73.

41. Archbald-Pannone LR, McMurry TL, Guerrant RL, Warren CA. Delirium and other clinical factors with Clostridium difficile infection that predict mortality in hospitalized patients. American journal of infection control. 2015;43(7):690–3.

42. Atamna A, Yahav D, Eliakim-Raz N, Goldberg E, Ben-Zvi H, Barsheshet A, et al. The effect of statins on the outcome of Clostridium difficile infection in hospitalized patients. European Journal of Clinical Microbiology and Infectious Diseases. 2016;35(5):779–84.

43. Barker AK, Van Galen A, Sethi AK, Shirley D, Safdar N. Tobacco use as a screener for Clostridium difficile infection outcomes. The Journal of hospital infection. 2018;98(1):36–9.

44. Bauer KA, Johnston JEW, Wenzler E, Goff DA, Cook CH, Balada-Llasat JM, et al. Impact of the NAP-1 strain on disease severity, mortality, and recurrence of healthcare-associated Clostridium difficile infection. Anaerobe. 2017;48:1–6.

45. Berry CE, Davies KA, Owens DW, Wilcox MH. Is there a relationship between the presence of the binary toxin genes in Clostridium difficile strains and the severity of C. difficile infection (CDI)? European journal of clinical microbiology & infectious diseases: official publication of the European Society of Clinical Microbiology. 2017;36(12):2405–15.

46. Boone JH, Archbald-Pannone LR, Wickham KN, Carman RJ, Guerrant RL, Franck CT, et al. Ribotype 027 Clostridium difficile infections with measurable stool toxin have increased lactoferrin and are associated with a higher mortality. European journal of clinical microbiology & infectious diseases: official publication of the European Society of Clinical Microbiology. 2014;33(6):1045–51.

47. Caupenne A, Ingrand P, Ingrand I, Forestier E, Roubaud-Baudron C, Gavazzi G, et al. Acute Clostridioides difficile Infection in Hospitalized Persons Aged 75 and Older: 30-Day Prognosis and Risk Factors for Mortality. Journal of the American Medical Directors Association. 2019.

48. Charilaou P, Devani K, John F, Kanna S, Ahlawat S, Young M, et al. Acute kidney injury impact on inpatient mortality in Clostridium difficile infection: A national propensity-matched study. Journal of gastroenterology and hepatology. 2018;33(6):1227–33.

49. Chintanaboina J, Navabi S, Suchniak-Mussari K, Stern B, Bedi S, Lehman EB, et al. Predictors of 30-Day Mortality in Hospitalized Patients with Clostridium difficile Infection. SouthMedJ. 2017;110(8):546–9.

50. Clohessy P, Merif J, Post JJ. Severity and frequency of community-onset Clostridium difficile infection on an Australian tertiary referral hospital campus. International Journal of Infectious Diseases. 2014;29:152–5.

51. Cohen NA, Miller T, Na’aminh W, Hod K, Adler A, Cohen D, et al. Clostridium difficile fecal toxin level is associated with disease severity and prognosis. United European gastroenterology journal. 2018;6(5):773–80.

52. Das R, Feuerstadt P, Brandt LJ. Glucocorticoids are associated with increased risk of short-term mortality in hospitalized patients with clostridium difficile-associated disease. Am J Gastroenterol. 2010;105(9):2040–9.

53. De Francesco MA, Lorenzin G, Piccinelli G, Corbellini S, Bonfanti C, Caruso A. Correlation between tcdB gene PCR cycle threshold and severe Clostridium difficile disease. Anaerobe. 2019;59:141–4.

54. Dudukgian H, Sie E, Gonzalez-Ruiz C, Etzioni DA, Kaiser AM. C. difficile colitis--predictors of fatal outcome. Journal of gastrointestinal surgery: official journal of the Society for Surgery of the Alimentary Tract. 2010;14(2):315–22.

55. Figh ML, Zoog ESL, Moore RA, Dart BWt, Heath G, Butler RM, et al. External Validation of Velazquez-Gomez Severity Score Index and ATLAS Scores and the Identification of Risk Factors Associated with Mortality in Clostridium difficile Infections. The American surgeon. 2017;83(12):1347–51.

56. Fountain EM, Moses MC, Park LP, Woods CW, Arepally GM. Thrombocytopenia in hospitalized patients with severe clostridium difficile infection. Journal of thrombosis and thrombolysis. 2017;43(1):38–42.

57. Fujii L, Fasolino J, Crowell MD, Dibaise JK. Appendectomy and risk of clostridium difficile recurrence. Infectious Diseases in Clinical Practice. 2013;21(1):28–32.

58. Hanania A, Jiang ZD, Smiley C, Lasco T, Garey KW, DuPont HL. Fecal calprotectin in the diagnosis of clostridium difficile infection. Infectious Diseases in Clinical Practice. 2016;24(1):31–4.

59. Henrich TJ, Krakower D, Bitton A, Yokoe DS. Clinical risk factors for severe Clostridium difficile-associated disease. Emerging infectious diseases. 2009;15(3):415–22.

60. Hubert B, Loo VG, Bourgault AM, Poirier L, Dascal A, Fortin E, et al. A portrait of the geographic dissemination of the Clostridium difficile North American pulsed-field type 1 strain and the epidemiology of C. difficile-associated disease in Quebec. Clin Infect Dis. 2007;44(2):238–44.

61. Huttunen R, Vuento R, Syrjanen J, Tissari P, Aittoniemi J. Case fatality associated with a hypervirulent strain in patients with culture-positive Clostridium difficile infection: a retrospective population-based study. International journal of infectious diseases: IJID: official publication of the International Society for Infectious Diseases. 2012;16(7):e532–5.

62. Kassam Z, Cribb Fabersunne C, Smith MB, Alm EJ, Kaplan GG, Nguyen GC, et al. Clostridium difficile associated risk of death score (CARDS): a novel severity score to predict mortality among hospitalised patients with C. difficile infection. Alimentary pharmacology & therapeutics. 2016;43(6):725–33.

63. Kenneally C, Rosini JM, Skrupky LP, Doherty JA, Hollands JM, Martinez E, et al. Analysis of 30-day mortality for clostridium difficile-associated disease in the ICU setting. Chest. 2007;132(2):418–24.

64. Khanafer N, Barbut F, Eckert C, Perraud M, Demont C, Luxemburger C, et al. Factors predictive of severe Clostridium difficile infection depend on the definition used. Anaerobe. 2016;37:43–8.

65. Khanna S, Pardi DS, Aronson SL, Kammer PP, Orenstein R, St Sauver JL, et al. The epidemiology of community-acquired Clostridium difficile infection: a population-based study. Am J Gastroenterol. 2012;107(1):89–95.

66. Khanna S, Gupta A, Baddour LM, Pardi DS. Epidemiology, outcomes, and predictors of mortality in hospitalized adults with Clostridium difficile infection. Internal and emergency medicine. 2016;11(5):657–65.

67. Khanna S, Keddis MT, Noheria A, Baddour LM, Pardi DS. Acute kidney injury is an independent marker of severity in Clostridium difficile infection: a nationwide survey. Journal of clinical gastroenterology. 2013;47(6):481–4.

68. Kim J, Kim H, Oh HJ, Kim HS, Hwang YJ, Yong D, et al. Fecal Calprotectin Level Reflects the Severity of Clostridium difficile Infection. Annals of laboratory medicine. 2017;37(1):53–7.

69. Kim J, Kim Y, Pai H. Clinical Characteristics and Treatment Outcomes of Clostridium difficile Infections by PCR Ribotype 017 and 018 Strains. PloS one. 2016;11(12):e0168849.

70. Kulaylat AS, Buonomo EL, Scully KW, Hollenbeak CS, Cook H, Petri WA, Jr., et al. Development and Validation of a Prediction Model for Mortality and Adverse Outcomes Among Patients With Peripheral Eosinopenia on Admission for Clostridium difficile Infection. JAMA surgery. 2018;153(12):1127–33.

71. Lee HC, Kim KO, Jeong YH, Lee SH, Jang BI, Kim TN. Clinical Outcomes in Hospitalized Patients with Clostridium difficile Infection by Age Group. The Korean journal of gastroenterology = Taehan Sohwagi Hakhoe chi. 2016;67(2):81–6.

72. Leibovici-Weissman Y, Atamna A, Schlesinger A, Eliakim-Raz N, Bishara J, Yahav D. Risk factors for short-and long-term mortality in very old patients with Clostridium difficile infection: A retrospective study. Geriatrics & gerontology international. 2017;17(10):1378–83.

73. Mascart G, Delmee M, Van Broeck J, Cytryn E, Karmali R, Cherifi S. Impact of ribotype 027 on Clostridium difficile infection in a geriatric department. European journal of clinical microbiology & infectious diseases: official publication of the European Society of Clinical Microbiology. 2013;32(9):1177–82.

74. Miller M, Gravel D, Mulvey M, Taylor G, Boyd D, Simor A, et al. Health care-associated Clostridium difficile infection in Canada: patient age and infecting strain type are highly predictive of severe outcome and mortality. Clin Infect Dis. 2010;50(2):194–201.

75. Morrison RH, Hall NS, Said M, Rice T, Groff H, Brodine SK, et al. Risk factors associated with complications and mortality in patients with Clostridium difficile infection. Clin Infect Dis. 2011;53(12):1173–8.

76. Ogielska M, Lanotte P, Le Brun C, Valentin AS, Garot D, Tellier AC, et al. Emergence of community-acquired Clostridium difficile infection: the experience of a French hospital and review of the literature. International journal of infectious diseases: IJID: official publication of the International Society for Infectious Diseases. 2015;37:36–41.

77. Patel UC, Wieczorkiewicz JT, Tuazon J. Evaluation of advanced age as a risk factor for severe Clostridium difficile infection. Journal of Clinical Gerontology and Geriatrics. 2016;7(1):12–6.

78. Pepin J, Valiquette L, Alary ME, Villemure P, Pelletier A, Forget K, et al. Clostridium difficile-associated diarrhea in a region of Quebec from 1991 to 2003: a changing pattern of disease severity. Cmaj. 2004;171(5):466–72.

79. Polivkova S, Krutova M, Petrlova K, Benes J, Nyc O. Clostridium difficile ribotype 176 - A predictor for high mortality and risk of nosocomial spread? Anaerobe. 2016;40:35–40.

80. Rao K, Micic D, Chenoweth E, Deng L, Galecki AT, Ring C, et al. Poor functional status as a risk factor for severe Clostridium difficile infection in hospitalized older adults. Journal of the American Geriatrics Society. 2013;61(10):1738–42.

81. Rao K, Micic D, Natarajan M, Winters S, Kiel MJ, Walk ST, et al. Clostridium difficile ribotype 027: relationship to age, detectability of toxins A or B in stool with rapid testing, severe infection, and mortality. Clin Infect Dis. 2015;61(2):233–41.

82. Reigadas E, Alcala L, Marin M, Martin A, Iglesias C, Bouza E. Role of binary toxin in the outcome of Clostridium difficile infection in a non-027 ribotype setting. Epidemiology and infection. 2016;144(2):268–73.

83. Sailhamer EA, Carson K, Chang Y, Zacharias N, Spaniolas K, Tabbara M, et al. Fulminant Clostridium difficile colitis: patterns of care and predictors of mortality. Arch Surg. 2009;144(5):433–9; discussion 9-40.

84. Scardina T, Labuszewski L, Pacheco SM, Adams W, Schreckenberger P, Johnson S. Clostridium difficile infection (CDI) severity and outcome among patients infected with the NAP1/BI/027 strain in a non-epidemic setting. Infect Control Hosp Epidemiol. 2015;36(3):280–6.

85. See I, Mu Y, Cohen J, Beldavs ZG, Winston LG, Dumyati G, et al. NAP1 strain type predicts outcomes from Clostridium difficile infection. Clin Infect Dis. 2014;58(10):1394–400.

86. Serafino S, Consonni D, Migone De Amicis M, Sisto F, Domeniconi G, Formica S, et al. Clinical outcomes of Clostridium difficile infection according to strain type. A prospective study in medical wards. European journal of internal medicine. 2018;54:21–6.

87. Shivashankar R, Khanna S, Kammer PP, Harmsen WS, Zinsmeister AR, Baddour LM, et al. Clinical factors associated with development of severe-complicated Clostridium difficile infection. Clinical gastroenterology and hepatology: the official clinical practice journal of the American Gastroenterological Association. 2013;11(11):1466–71.

88. Starzengruber P, Segagni Lusignani L, Wrba T, Mitteregger D, Indra A, Graninger W, et al. Severe Clostridium difficile infection: incidence and risk factors at a tertiary care university hospital in Vienna, Austria. Wiener klinische Wochenschrift. 2014;126(13-14):427–30.

89. Tamez-Torres KM, Torres-Gonzalez P, Leal-Vega F, Garcia-Alderete A, Lopez Garcia NI, Mendoza-Aguilar R, et al. Impact of Clostridium difficile infection caused by the NAP1/RT027 strain on severity and recurrence during an outbreak and transition to endemicity in a Mexican tertiary care center. International journal of infectious diseases: IJID: official publication of the International Society for Infectious Diseases. 2017;65:44–9.

90. Taori SK, Wroe A, Hardie A, Gibb AP, Poxton IR. A prospective study of community-associated Clostridium difficile infections: the role of antibiotics and co-infections. The Journal of infection. 2014;69(2):134–44.

91. Tschudin-Sutter S, Braissant O, Erb S, Stranden A, Bonkat G, Frei R, et al. Growth Patterns of Clostridium difficile - Correlations with Strains, Binary Toxin and Disease Severity: A Prospective Cohort Study. PloS one. 2016;11(9):e0161711.

92. van der Wilden GM, Chang Y, Cropano C, Subramanian M, Schipper IB, Yeh DD, et al. Fulminant Clostridium difficile colitis: prospective development of a risk scoring system. The journal of trauma and acute care surgery. 2014;76(2):424–30.

93. Vojtilova L, Freibergerova M, Jurankova J, Bortlicek Z, Husa P. Epidemiological factors influencing the development of relapsing and severe Clostridium difficile infection. Epidemiologie, mikrobiologie, imunologie: casopis Spolecnosti pro epidemiologii a mikrobiologii Ceske lekarske spolecnosti JE Purkyne. 2014;63(1):27–35.

94. Walk ST, Micic D, Jain R, Lo ES, Trivedi I, Liu EW, et al. Clostridium difficile Ribotype Does Not Predict Severe Infection. Clin Infect Dis. 2012;55(12):1661–8.

95. Walker AS, Eyre DW, Wyllie DH, Dingle KE, Griffiths D, Shine B, et al. Relationship between bacterial strain type, host biomarkers, and mortality in Clostridium difficile infection. Clin Infect Dis. 2013;56(11):1589–600.

96. Wilson V, Cheek L, Satta G, Walker-Bone K, Cubbon M, Citron D, et al. Predictors of death after Clostridium difficile infection: a report on 128 strain-typed cases from a teaching hospital in the United Kingdom. Clin Infect Dis. 2010;50(12):e77–81.

97. Xu Q, Chen Y, Gu S, Lv T, Zheng B, Shen P, et al. Hospital-acquired Clostridium difficile infection in Mainland China: A seven-year (2009-2016) retrospective study in a large university hospital. Scientific reports. 2017;7(1):9645.

98. Yong FA, Alvarado AM, Wang H, Tsai J, Estes NC. Appendectomy: a risk factor for colectomy in patients with Clostridium difficile. American journal of surgery. 2015;209(3):532–5.

99. Miller R, Morillas JA, Brizendine KD, Fraser TG. Predictors of Clostridioides difficile Infection-Related Complications and Treatment Patterns among Nucleic Acid Amplification Test-Positive/Toxin Enzyme Immunoassay-Negative Patients. J Clin Microbiol. 2020;58(3).

100. Menon A, Perry DA, Motyka J, Weiner S, Standke A, Penkevich A, et al. Changes in the Association between Diagnostic Testing Method, PCR Ribotype, and Clinical Outcomes from Clostridioides difficile Infection: One Institution’s Experience. Clin Infect Dis. 2020.

101. Korac M, Rupnik M, Nikolic N, Jovanovic M, Tosic T, Malinic J, et al. Clostridioides difficile ribotype distribution in a large teaching hospital in Serbia. Gut Pathog. 2020;12:26.

102. Essrani R, Saturno D, Mehershahi S, Essrani RK, Hossain MR, Ravi SJK, et al. The Impact of Appendectomy in Clostridium difficile Infection and Length of Hospital Stay. Cureus. 2020;12(9):e10342.

103. Choi B, Wong KK, Dunn AN, Butler R, Fraser TG, Procop GW, et al. Real-time polymerase chain reaction (PCR) cycle threshold and Clostridioides difficile infection outcomes. Infect Control Hosp Epidemiol. 2021:1–7.

104. Avni T, Hammud H, Itzhaki O, Gafter-Gvili A, Rozen-Zvi B, Ben-Zvi H, et al. The significance of acute kidney injury in Clostridioides difficile infection. Int J Clin Pract. 2021;75(3).

105. Mendez-Bailon M, Jimenez-Garcia R, Hernandez-Barrera V, Miguel-Diez J, Miguel-Yanes JM, Munoz-Rivas N, et al. Heart Failure Is a Risk Factor for Suffering and Dying of Clostridium difficile Infection. Results of a 15-Year Nationwide Study in Spain. J Clin Med. 2020;9(3).

106. Chiang HY, Huang HC, Chung CW, Yeh YC, Chen YC, Tien N, et al. Risk prediction for 30-day mortality among patients with Clostridium difficile infections: a retrospective cohort study. Antimicrob Resist Infect Control. 2019;8:175.

107. Vata A, Miftode IL, Dorneanu OS, Vata LG, Miftode D, Trifan A, et al. CLOSTRIDIUM DIFFICILE INFECTION IN PATIENTS WITH TYPE 2 DIABETES MELLITUS. THE EXPERIENCE OF AN INFECTIOUS DISEASES HOSPITAL FROM NORTH-EASTERN ROMANIA. Med-Surg J. 2019;123(3):506–12.

108. Mani NS, Lynch JB, Fang FC, Chan JD. Risk Factors for BI/NAP1/027 Clostridioides difficile Infections and Clinical Outcomes Compared With Non-NAP1 Strains. Open Forum Infect Dis. 2019;6(12):4.

109. Khanna S, Aronson SL, Kammer PP, Baddour LM, Pardi DS. Gastric acid suppression and outcomes in Clostridium difficile infection: a population-based study. Mayo Clinic proceedings. 2012;87(7):636–42.

110. Planche TD, Davies KA, Coen PG, Finney JM, Monahan IM, Morris KA, et al. Differences in outcome according to Clostridium difficile testing method: a prospective multicentre diagnostic validation study of C difficile infection. The Lancet Infectious diseases. 2013;13(11):936–45.

111. Bauer MP, Notermans DW, van Benthem BH, Brazier JS, Wilcox MH, Rupnik M, et al. Clostridium difficile infection in Europe: a hospital-based survey. Lancet. 2011;377(9759):63–73.

112. Argamany JR, Lee GC, Duhon BD, Zeidan AR, Young EH, Reveles KR. A possible association between statin use and improved Clostridioides difficile infection mortality in veterans. PloS one. 2019;14(5):e0217423.

113. Leal J, Ronksley P, Henderson EA, Conly J, Manns B. Predictors of mortality and length of stay in patients with hospital-acquired Clostridioides difficile infection: a population-based study in Alberta, Canada. The Journal of hospital infection. 2019;103(1):85–91.

114. Nagayoshi Y, Yamamoto K, Sato S, Suyama N, Izumikawa T, Izumikawa K, et al. Clinical significance of a positive Clostridioides difficile glutamate dehydrogenase test on the outcomes of hospitalized older patients. Geriatrics & gerontology international. 2020;20(12):1138–44.

115. Patel H, Makker J, Vakde T, Shaikh D, Badipatla K, Dunne J, et al. Nonsteroidal Anti-Inflammatory Drugs Impact on the Outcomes of Hospitalized Patients with Clostridium difficile Infection. Clinical and experimental gastroenterology. 2019;12:449–56.

116. Tay HL, Chow A, Ng TM, Lye DC. Risk factors and treatment outcomes of severe Clostridioides difficile infection in Singapore. Scientific reports. 2019;9(1):13440.

117. Origuen J, Orellana MA, Fernandez-Ruiz M, Corbella L, San Juan R, Ruiz-Ruigomez M, et al. Toxin B PCR Amplification Cycle Threshold Adds Little to Clinical Variables for Predicting Outcomes in Clostridium difficile Infection: a Retrospective Cohort Study. J Clin Microbiol. 2019;57(2).

118. Milenkovic B, Suljagic V, Peric A, Dragojevic-Simic V, Tarabar O, Milanovic M, et al. Outcomes of Clostridioides difficile infection in adult cancer and non-cancer patients hospitalised in a tertiary hospital: a prospective cohort study. Eur J Hosp Pharm. 2021.

119. Lopez-Cardenas S, Torres-Martos E, Mora-Delgado J, Sanchez-Calvo JM, Santos-Pena M, Zapata Lopez A, et al. The prognostic value of toxin B and binary toxin in Clostridioides difficile infection. Gut Microbes. 2021:1–8.

120. Enoch DA, Murray-Thomas T, Adomakoh N, Dedman D, Georgopali A, Francis NA, et al. Risk of complications and mortality following recurrent and non-recurrent Clostridioides difficile infection: a retrospective observational database study in England. The Journal of hospital infection. 2020;106(4):793–803.

121. Carlson TJ, Endres BT, Le Pham J, Gonzales-Luna AJ, Alnezary FS, Nebo K, et al. Eosinopenia and Binary Toxin Increase Mortality in Hospitalized Patients With Clostridioides difficile Infection. Open Forum Infect Dis. 2020;7(1):ofz552.

122. Abdelfatah M, Nayfe R, Nijim A, Enriquez K, Ali E, Watkins RR, et al. Factors Predicting Recurrence of Clostridium difficile Infection (CDI) in Hospitalized Patients: Retrospective Study of More Than 2000 Patients. J Investig Med. 2015;63(5):747–51.

123. Appaneal HJ, Caffrey AR, Beganovic M, Avramovic S, LaPlante KL. Predictors of Clostridioides difficile recurrence across a national cohort of veterans in outpatient, acute, and long-term care settings. Am J Health Syst Pharm. 2019;76(9):581–90.

124. Avni T, Babitch T, Ben-Zvi H, Hijazi R, Ayada G, Atamna A, et al. Clostridioides difficile infection in immunocompromised hospitalized patients is associated with a high recurrence rate. Int J Infect Dis. 2020;90:237–42.

125. Carpenter BP, Hennessey EK, Bryant AM, Khoury JA, Crannage AJ. Identification of Factors Impacting Recurrent Clostridium difficile Infection and Development of a Risk Evaluation Tool. J Pharm Pharm Sci. 2016;19(3):349–56.

126. Dharbhamulla N, Abdelhady A, Domadia M, Patel S, Gaughan J, Roy S. Risk Factors Associated With Recurrent Clostridium difficile Infection. J Clin Med Res. 2019;11(1):1–6.

127. Drekonja DM, Amundson WH, Decarolis DD, Kuskowski MA, Lederle FA, Johnson JR. Antimicrobial use and risk for recurrent Clostridium difficile infection. Am J Med. 2011;124(11):1081 e1-7.

128. Eyre DW, Walker AS, Wyllie D, Dingle KE, Griffiths D, Finney J, et al. Predictors of first recurrence of Clostridium difficile infection: implications for initial management. Clin Infect Dis. 2012;55 Suppl 2:S77–87.

129. Falcone M, Tiseo G, Iraci F, Raponi G, Goldoni P, Delle Rose D, et al. Risk factors for recurrence in patients with Clostridium difficile infection due to 027 and non-027 ribotypes. Clin Microbiol Infect. 2019;25(4):474–80.

130. Freedberg DE, Salmasian H, Friedman C, Abrams JA. Proton pump inhibitors and risk for recurrent Clostridium difficile infection among inpatients. Am J Gastroenterol. 2013;108(11):1794–801.

131. Fujii LF, J. Crowell, M. DiBaise K. Appendectomy and Risk of Clostridium difficile Recurrence. Infectious Diseases in Clinical Practice. 2013;21(1):28–32.

132. Im GY, Modayil RJ, Lin CT, Geier SJ, Katz DS, Feuerman M, et al. The appendix may protect against Clostridium difficile recurrence. Clinical gastroenterology and hepatology: the official clinical practice journal of the American Gastroenterological Association. 2011;9(12):1072–7.

133. Kimura T, Snijder R, Sugitani T. Characterization and risk factors for recurrence of Clostridioides (Clostridium) difficile infection in Japan: A nationwide real-world analysis using a large hospital-based administrative dataset. J Infect Chemother. 2019;25(8):615–20.

134. Lavergne V, Beausejour Y, Pichette G, Ghannoum M, Su SH. Lymphopenia as a novel marker of Clostridium difficile infection recurrence. The Journal of infection. 2013;66(2):129–35.

135. Linsky A, Gupta K, Lawler EV, Fonda JR, Hermos JA. Proton pump inhibitors and risk for recurrent Clostridium difficile infection. Arch Intern Med. 2010;170(9):772–8.

136. Louie TJ, Miller MA, Crook DW, Lentnek A, Bernard L, High KP, et al. Effect of age on treatment outcomes in Clostridium difficile infection. Journal of the American Geriatrics Society. 2013;61(2):222–30.

137. Lupse M, Flonta M, Cioara A, Filipescu I, Todor N. Predictors of first recurrence in Clostridium difficile-associated disease. A study of 306 patients hospitalized in a Romanian tertiary referral center. J Gastrointestin Liver Dis. 2013;22(4):397–403.

138. Marincu I, Bratosin F, Vidican I, Cerbu B, Turaiche M, Tirnea L, et al. Predictive Factors for the First Recurrence of Clostridioides difficile Infection in the Elderly from Western Romania. Medicina (Kaunas). 2020;56(9).

139. McDonald EG, Milligan J, Frenette C, Lee TC. Continuous Proton Pump Inhibitor Therapy and the Associated Risk of Recurrent Clostridium difficile Infection. JAMA Intern Med. 2015;175(5):784–91.

140. McFarland LV, Surawicz CM, Rubin M, Fekety R, Elmer GW, Greenberg RN. Recurrent Clostridium difficile disease: epidemiology and clinical characteristics. Infect Control Hosp Epidemiol. 1999;20(1):43–50.

141. Na’amnih W, Adler A, Miller-Roll T, Cohen D, Carmeli Y. Risk factors for recurrent Clostridium difficile infection in a tertiary hospital in Israel. European journal of clinical microbiology & infectious diseases: official publication of the European Society of Clinical Microbiology. 2018;37(7):1281–8.

142. Negrut N, Bungau S, Behl T, Khan SA, Vesa CM, Bustea C, et al. Risk Factors Associated with Recurrent Clostridioides difficile Infection. Healthcare. 2020;8(3):12.

143. Origüen J, Orellana M, Fernández-Ruiz M, Corbella L, San Juan R, Ruiz-Ruigómez M, et al. Toxin B PCR Amplification Cycle Threshold Adds Little to Clinical Variables for Predicting Outcomes in Clostridium difficile Infection: a Retrospective Cohort Study. J Clin Microbiol. 2019;57(2).

144. Pepin J, Alary ME, Valiquette L, Raiche E, Ruel J, Fulop K, et al. Increasing risk of relapse after treatment of Clostridium difficile colitis in Quebec, Canada. Clin Infect Dis. 2005;40(11):1591–7.

145. Rao K, Higgins PDR, Young VB. An Observational Cohort Study of Clostridium difficile Ribotype 027 and Recurrent Infection. mSphere. 2018;3(3).

146. Rodriguez-Pardo D, Almirante B, Bartolome RM, Pomar V, Mirelis B, Navarro F, et al. Epidemiology of Clostridium difficile infection and risk factors for unfavorable clinical outcomes: results of a hospital-based study in Barcelona, Spain. J Clin Microbiol. 2013;51(5):1465–73.

147. Rotramel A, Poritz LS, Messaris E, Berg A, Stewart DB. PPI therapy and albumin are better predictors of recurrent Clostridium difficile colitis than choice of antibiotics. ournal of gastrointestinal surgery: official journal of the Society for Surgery of the Alimentary Tract. 2012;16(12):2267–73.

148. van Beurden YH, Nezami S, Mulder CJJ, Vandenbroucke-Grauls C. Host factors are more important in predicting recurrent Clostridium difficile infection than ribotype and use of antibiotics. Clin Microbiol Infect. 2018;24(1):85 e1-e4.

149. Zilberberg MD, Reske K, Olsen M, Yan Y, Dubberke ER. Risk factors for recurrent Clostridium difficile infection (CDI) hospitalization among hospitalized patients with an initial CDI episode: a retrospective cohort study. BMC Infect Dis. 2014;14:306.

150. Cadena J, Thompson GR, 3rd, Patterson JE, Nakashima B, Owens A, Echevarria K, et al. Clinical predictors and risk factors for relapsing Clostridium difficile infection. Am J Med Sci. 2010;339(4):350–5.

151. Shakov R, Salazar RS, Kagunye SK, Baddoura WJ, DeBari VA. Diabetes mellitus as a risk factor for recurrence of Clostridium difficile infection in the acute care hospital setting. American journal of infection control. 2011;39(3):194–8.

152. Fekety R, McFarland LV, Surawicz CM, Greenberg RN, Elmer GW, Mulligan ME. Recurrent Clostridium difficile diarrhea: characteristics of and risk factors for patients enrolled in a prospective, randomized, double-blinded trial. Clin Infect Dis. 1997;24(3):324–33.

153. Ryu HS, Kim YS, Seo GS, Lee YM, Choi SC. Risk Factors for Recurrent Clostridium difficile Infection. Intest Res. 2012;10(2):176–82.

154. Golan Y, DuPont HL, Aldomiro F, Jensen EH, Hanson ME, Dorr MB. Renal Impairment, C. difficile Recurrence, and the Differential Effect of Bezlotoxumab: A Post Hoc Analysis of Pooled Data From 2 Randomized Clinical Trials. Open Forum Infect Dis. 2020;7(7):ofaa248.

155. Weiss K, Louie T, Miller MA, Mullane K, Crook DW, Gorbach SL. Effects of proton pump inhibitors and histamine-2 receptor antagonists on response to fidaxomicin or vancomycin in patients with Clostridium difficile-associated diarrhoea. BMJ Open Gastroenterol. 2015;2(1):e000028.

156. Gerding DN, Kelly CP, Rahav G, Lee C, Dubberke ER, Kumar PN, et al. Bezlotoxumab for Prevention of Recurrent Clostridium difficile Infection in Patients at Increased Risk for Recurrence. Clin Infect Dis. 2018;67(5):649–56.

157. Crook DW, Walker AS, Kean Y, Weiss K, Cornely OA, Miller MA, et al. Fidaxomicin versus vancomycin for Clostridium difficile infection: meta-analysis of pivotal randomized controlled trials. Clin Infect Dis. 2012;55 Suppl 2:S93–103.

158. Wilcox MH, Gerding DN, Poxton IR, Kelly C, Nathan R, Birch T, et al. Bezlotoxumab for Prevention of Recurrent Clostridium difficile Infection. N Engl J Med. 2017;376(4):305–17.

159. Louie TJ, Miller MA, Mullane KM, Weiss K, Lentnek A, Golan Y, et al. Fidaxomicin versus vancomycin for Clostridium difficile infection. N Engl J Med. 2011;364(5):422–31.

160. Cornely OA, Crook DW, Esposito R, Poirier A, Somero MS, Weiss K, et al. Fidaxomicin versus vancomycin for infection with Clostridium difficile in Europe, Canada, and the USA: a double-blind, non-inferiority, randomised controlled trial. The Lancet Infectious diseases. 2012;12(4):281–9.

161. Charlson ME, Pompei P, Ales KL, MacKenzie CR. A new method of classifying prognostic comorbidity in longitudinal studies: development and validation. Journal of chronic diseases. 1987;40(5):373–83.

162. Abou Chakra CN, Pepin J, Sirard S, Valiquette L. Risk factors for recurrence, complications and mortality in Clostridium difficile infection: a systematic review. PloS one. 2014;9(6):e98400.

163. Deshpande A, Pasupuleti V, Thota P, Pant C, Rolston DD, Hernandez AV, et al. Risk factors for recurrent Clostridium difficile infection: a systematic review and meta-analysis. Infect Control Hosp Epidemiol. 2015;36(4):452–60.

164. Garey KW, Sethi S, Yadav Y, DuPont HL. Meta-analysis to assess risk factors for recurrent Clostridium difficile infection. The Journal of hospital infection. 2008;70(4):298–304.

165. Zhang VRY, Woo ASJ, Scaduto C, Cruz MTK, Tan YY, D. H, et al. Systematic review on the definition and predictors of severe Clostridiodes difficile infection. Journal of gastroenterology and hepatology. 2021;36(1):89–104.

166. Dahabreh IJ, Kent DM. Index event bias as an explanation for the paradoxes of recurrence risk research. Jama. 2011;305(8):822–3.

167. Phatharacharukul P, Thongprayoon C, Cheungpasitporn W, Edmonds PJ, Mahaparn P, Bruminhent J. The Risks of Incident and Recurrent Clostridium difficile-Associated Diarrhea in Chronic Kidney Disease and End-Stage Kidney Disease Patients: A Systematic Review and Meta-Analysis. Digestive diseases and sciences. 2015;60(10):2913–22.

168. Dalal RS, Allegretti JR. Diagnosis and management of Clostridioides difficile infection in patients with inflammatory bowel disease. Current opinion in gastroenterology. 2021;37(4):336–43.

169. Beauregard-Paultre C, Abou Chakra CN, McGeer A, Labbé AC, Simor AE, Gold W, et al. External validation of clinical prediction rules for complications and mortality following Clostridioides difficile infection. PloS one. 2019;14(12):e0226672.

170. van Beurden YH, Hensgens MPM, Dekkers OM, Le Cessie S, Mulder CJJ, Vandenbroucke-Grauls C. External Validation of Three Prediction Tools for Patients at Risk of a Complicated Course of Clostridium difficile Infection: Disappointing in an Outbreak Setting. Infect Control Hosp Epidemiol. 2017;38(8):897–905.

171. van Rossen TM, van Dijk LJ, Heymans MW, Dekkers OM, Vandenbroucke-Grauls C, van Beurden YH. External validation of two prediction tools for patients at risk for recurrent Clostridioides difficile infection. Therapeutic advances in gastroenterology. 2021;14:1756284820977385.

